# Prevalence of non-communicable diseases risk factors and their determinants in Malawi: Evidence from 2017 WHO STEPwise Survey

**DOI:** 10.1101/2022.08.18.22278928

**Authors:** Wingston Felix Ng’ambi, Takondwa Mwase, Jobiba Chinkhumba, Michael Udedi, Farai Chigaru, Jonathan Chiwanda Banda, Dominic Nkhoma, Joseph Mfutso-Bengo

## Abstract

**Introduction:** By 2030, the non-communicable diseases (NCDs) are expected to overtake communicable, maternal, neonatal, and nutritional (CMNN) diseases combined as the leading cause of mortality in sub-Saharan Africa (SSA). With the increasing trend in NCDs, the NCD risk factors (NCDRF) need to be understood at local level in order to guide NCD risk mitigation efforts. Therefore, we provide a detailed analysis of some modifiable NCDRF and their determinants in Malawi using the 2017 Stepwise survey (STEPS).

**Methods:** This is a secondary analysis of the Malawi 2017 STEPS. Data was analysed using frequencies, proportions, odds ratios (OR) and their associated 95% confidence intervals (95%CI). We fitted multiple logistic regression of the NCD risk factors on the explanatory variables using likelihood ratio test. The level of statistical significance was set at P< 0.05.

**Results:** Of the 4187 persons, 9% were current smokers, 1% were taking alcohol, 16% had high salt intake, 64% had insufficient fruit intake, 21% had low physical activity, 25% had high blood sugar, and 11% had high blood pressure. Smoking odds increased with age but decreased with level of education. Females had lower odds of engaging in harmful alcohol use than males (AOR=0.04, 95%CI: 0.01-0.17, P<0.001). Females had lower odds of high salt uptake than the males (AOR=0.70, 95%CI: 0.58-0.84, P=0.0001). Persons in non-paid jobs had higher odds of salt uptake than those employed (AOR=1.70, 95%CI: 1.03-2.79, P=0.04). Females were 22% more likely to have insufficient fruit uptake compared to males (AOR=1.22, 95%CI: 1.06-1.41, P=0.007).

**Conclusion:** The high prevalence of physical inactivity, high salt consumption, insufficient fruit intake, raised blood glucose and high relatively blood pressure calls for a sound public health approach. The Malawi Ministry of Health should devise multi-sectoral approaches that minimize exposure to modifiable NCD risk factors at population and individual levels.

## INTRODUCTION

There is an increase in mortality from non-communicable diseases (NCDs) globally with nearly 75% of these deaths occurring in low-middle income countries (LMIs) [1]. The sub-Saharan Africa (SSA) is facing an epidemiological transition with the NCDs emerging as the leading causes of deaths [2]. There has been an increased burden of NCDs in sub-Saharan Africa over the past two decades, driven by increasing incidence of cardiovascular risk factors such as unhealthy diets, reduced physical activity, hypertension, obesity, diabetes, dyslipidaemia, and air pollution [3]. By 2030, the NCDs are expected to overtake communicable, maternal, neonatal, and nutritional (CMNN) diseases combined as the leading cause of mortality in sub-Saharan Africa (SSA) [4].

With the increasing trend in NCDs, the NCD risk factors (NCDRF) need to be understood at local level in order to guide NCD risk mitigation efforts [5]. Besides, the several studies done in Malawi on the NCD risk factors [2] [6] [7] [8] [9] [10] have only considered a few socio-demographic factors or have limited analysis of the determinants of the NCD risk factors. Therefore, we conducted this study in order to have a comprehensive understanding of the risk factors for NCDs and their determinants in Malawi using the 2017 Malawi National STEP Survey.

## METHODS

### Study design

This is a secondary analysis of the Malawi 2017 STEPS [11]. This analysis includes data from all the 28 districts of Malawi. The 2017 Malawi STEPS used the WHO STEPwise approach to assessing risk factors for chronic non-communicable diseases [7]. The Malawi 2017 STEP survey used a multi-stage cluster sample of households. One individual within the age range of the survey (18-69) was selected per household [10].

### Statistical analysis and data management

The data were managed in Stata version 17.0 (StataCorp, Texas, USA). The outcome variables comprised five behavioural (current tobacco use, harmful alcohol consumption, low consumption of fruits, physical inactivity, and high salt intake) and two biological (history of raised blood pressure and raised blood glucose) risk factors of NCD [12]. Age, education, occupation, residence, sex of respondent, and marital status were the explanatory variables adopted for this study.

Data was analysed using frequencies, proportions, odds ratios (OR) and their associated 95% confidence intervals (95%CI). A multivariate logistic regression model was used to quantify the determinants of NCDs risk factors with each risk factor as a dependent variable. We fitted a multiple logistic regression model of the NCD risk factors using a forward step-wise selection method. Age and sex were entered as a priori variables in the multiple logistic regression models. For a factor to be included in the model, likelihood ratio test (LRT) was used. Data was presented using tables and figures. The level of statistical significance was set at P< 0.05.

### Ethical consideration

We requested access for secondary use of the data from the World Health Organization, the funder for the Malawi 2017 STEPS survey. The Malawi 2017 STEPS survey dataset was downloaded from https://extranet.who.int/ncdsmicrodata/index.php/catalog/629.

## RESULTS

### Characteristics of respondents

The socio-demographic characteristics of the men and women assessed for NCD risk factors are shown in Table 1. In total 4187 persons were assessed for NCD risk factors. Of these, 65% (2702) were females; 28% (1164) were aged between 25 years and 34 years while 15% (617) were aged between 45 years and 54 years; 61% (2545) had primary school as their highest level of education attainment while 4% (148) had tertiary education as their highest level of education; 69% (2888) were currently married while 10% (418) were never married; and 35% (1451) were self-employed while 4% (150) were employed by the Government of Malawi. Missing data ranged from <1% (15 of 4187) to 2% (150 of 4187)

**Table 1:** Characteristics of respondents that participated in Malawi STEPS surveys: 2017

| Characteristics | Total |  |
| --- | --- | --- |
|  | n | % |
| **Total** | 4073 | 100.0 |
| **Residence** |  |  |
| *Rural* | 3247 | 79.7 |
| *Urban* | 826 | 20.3 |
| **Sex** |  |  |
| *Male* | 1485 | 35.5 |
| *Female* | 2702 | 64.5 |
| **Age group** |  |  |
| 15-24 | 788 | 18.8 |
| 25-34 | 1164 | 27.8 |
| 35-44 | 967 | 23.1 |
| 45-54 | 617 | 14.7 |
| 55+ | 651 | 15.5 |
| **Highest education level** |  |  |
| *No education* | 585 | 14.0 |
| *Primary* | 2545 | 60.8 |
| *Secondary* | 909 | 21.7 |
| *Tertiary* | 148 | 3.5 |
| **Marital status** |  |  |
| *Never married* | 418 | 10.0 |
| *Currently married* | 2888 | 69.0 |
| *Not Married* | 866 | 20.7 |
| *Missing* | 15 | 0.4 |
| **Occupation** |  |  |
| *Government employee* | 150 | 3.6 |
| *Non-Govt employee* | 155 | 3.7 |
| *Self-employed* | 1451 | 34.7 |
| *Non-paid* | 1362 | 32.5 |
| *Retired* | 967 | 23.1 |
| *Missing* | 102 | 2.4 |

### Prevalence of selected risk factors for NCDs

#### Tobacco use

The overall prevalence of current tobacco use was 9%. There was an increasing trend in tobacco use by age of an individual (see Table 2). Persons with no education had the highest tobacco use (11%) while those with tertiary education had the least use of tobacco (5%). The use of tobacco was highest amongst those who are currently married (9%) and least amongst those who have never been married (7%). The persons who retired had the least use of tobacco (4%) compared to those working in non-governmental organizations (14%). A higher proportion of men (21%) than women (2%) were using tobacco (see Figure 1).

**Table 2:** Prevalence of NCDs risk factors stratified by socio-demographic characteristics of respondents in Malawi: 2017 STEPS survey

| Characteristics (n=4073) | Current smoker | Harmful alcohol use | High salt intake | Low fruit intake | Physical inactivity | Raised blood sugar | Raised blood pressure |
| --- | --- | --- | --- | --- | --- | --- | --- |
| **Overall** | 8.60 (7.75-9.45) | 0.69 (0.44-0.94) | 16.36 (15.24-17.48) | 64.17 (62.72-65.63) | 21.18 (19.95-22.42) | 25.29 (23.28-27.30) | 10.69 (9.75-11.63) |
| **Residence** |  |  |  |  |  |  |  |
| *Rural* | 9.12 (8.13-10.11) | 0.68 (0.40-0.96) | 15.61 (14.37-16.86) | 60.98 (59.30-62.66) | 22.27 (20.84-23.70) | 9.06 (8.07-10.05) | 22.64 (20.34-24.94) |
| *Urban* | 6.30 (4.64-7.95) | 0.85 (0.22-1.45) | 21.07 (18.28-23.85) | 72.76 (69.72-75.80) | 19.61 (16.90-22.32) | 17.80 (15.19-20.41) | 32.01 (27.92-36.09) |
| **Age group** |  |  |  |  |  |  |  |
| *15-24* | 3.05 (1.84-4.25) | 0.51 (0.01-1.00) | 16.75 (14.14-19.36) | 71.57 (68.42-74.73) | 28.68 (25.52-31.84) | 11.90 (8.19-15.62) | 12.47 (10.16-14.78) |
| *25-34* | 6.79 (5.34-8.23) | 0.52 (0.10-0.93) | 18.30 (16.07-20.52) | 68.30 (65.62-70.97) | 25.43 (22.93-27.93) | 14.47 (11.53-17.40) | 11.90 (10.03-13.76) |
| *35-44* | 10.44 (8.52-12.37) | 1.24 (0.54-1.94) | 15.51 (13.23-17.80) | 63.81 (60.77-66.84) | 20.17 (17.63-22.70) | 25.50 (21.22-29.78) | 11.54 (9.52-13.56) |
| *45-54* | 11.51 (8.99-14.03) | 0.64 (0.01-1.28) | 15.88 (13.0-18.77) | 60.13 (56.26-64.00) | 14.91 (12.10-17.72) | 37.31 (31.41-43.20) | 7.29 (5.24-9.35) |
| *55+* | 13.06 (10.47-15.65) | 0.46 (-0.06-0.98) | 14.13 (11.45-16.81) | 52.23 (48.39-56.07) | 11.98 (9.48-14.48) | 47.97 (42.27-53.68) | 8.33 (6.20-10.46) |
| **Highest education level** |  |  |  |  |  |  |  |
| *No education* | 11.11 (8.56-13.66) | 0.17 (-0.16-0.51) | 15.56 (12.62-18.50) | 49.23 (45.17-53.29) | 8.72 (6.43-11.01) | 34.33 (28.22-40.45) | 8.46 (6.19-10.79) |
| *Primary* | 9.31 (8.18-10.44) | 0.71 (0.38-1.03) | 14.81 (13.43-16.19) | 63.03 (61.15-64.90) | 19.45 (17.91-20.99) | 24.47 (21.84-27.09) | 9.58 (8.43-10.72) |
| *Secondary* | 5.61 (4.11-7.11) | 0.77 (0.20-1.34) | 18.92 (16.37-21.47) | 73.82 (70.96-76.68) | 32.12 (29.09-35.16) | 23.86 (19.87-27.85) | 13.20 (11.00-15.40) |
| *Tertiary* | 4.73 (1.30-8.16) | 2.03 (-0.25-4.31) | 30.41 (22.97-37.84) | 83.78 (77.82-89.74) | 33.11 (25.50-40.72) | 19.00 (11.27-26.73) | 22.97 (16.17-29.78) |
| **Marital status** |  |  |  |  |  |  |  |
| *Never married* | 6.70 (4.30-9.10) | 1.20 (0.15-2.24) | 19.62 (15.80-23.43) | 68.66 (64.21-73.11) | 32.30 (27.81-36.79) | 21.82 (14.06-29.58) | 15.07 (11.64-18.51) |
| *Currently married* | 9.04 (7.99-10.08) | 0.69 (0.39-1.00) | 16.97 (15.60-18.34) | 64.54 (62.80-66.29) | 20.60 (19.13-22.08) | 24.06 (21.71-26.41) | 10.52 (9.40-11.64) |
| *Not Married* | 7.97 (6.16-9.77) | 0.46 (0.01-0.91) | 12.70 (10.48-14.92) | 60.74 (57.48-63.99) | 17.90 (15.34-20.45) | 29.95 (25.53-34.37) | 8.95 (7.04-10.86) |
| **Occupation** |  |  |  |  |  |  |  |
| *Government employee* | 5.33 (1.72-8.94) | 0.67 (-0.64-1.97) | 22.67 (15.94-29.39) | 72.67 (65.51-79.82) | 36.67 (28.93-44.41) | 25.00 (15.89-34.11) | 13.33 (2.78-7.87) |
| *Non-Govt employee* | 13.55 (8.14-18.96) | 0.65 (-0.62-1.91) | 19.35 (13.11-25.60) | 69.68 (62.42-76.94) | 26.45 (19.48-33.42) | 29.03 (17.63-40.43) | 13.55 (2.76-8.14) |
| *Self-employed* | 10.27 (8.71-11.83) | 1.17 (0.62-1.73) | 15.23 (13.38-17.08) | 67.88 (65.48-70.29) | 26.19 (23.93-28.45) | 21.13 (17.86-24.40) | 11.53 (0.84-9.88) |
| *Non-paid* | 9.99 (8.39-11.58) | 0.44 (0.09-0.79) | 21.66 (19.47-23.85) | 57.34 (54.71-59.97) | 20.93 (18.76-23.09) | 27.61 (23.82-31.40) | 10.65 (0.84-9.01) |
| *Retired* | 3.62 (2.44-4.80) | 0.41 (0.01-0.82) | 10.75 (8.80-12.71) | 62.56 (59.51-65.62) | 13.03 (10.91-15.15) | 27.49 (23.54-31.45) | 9.43 (0.94-7.58) |

**Figure 1:**
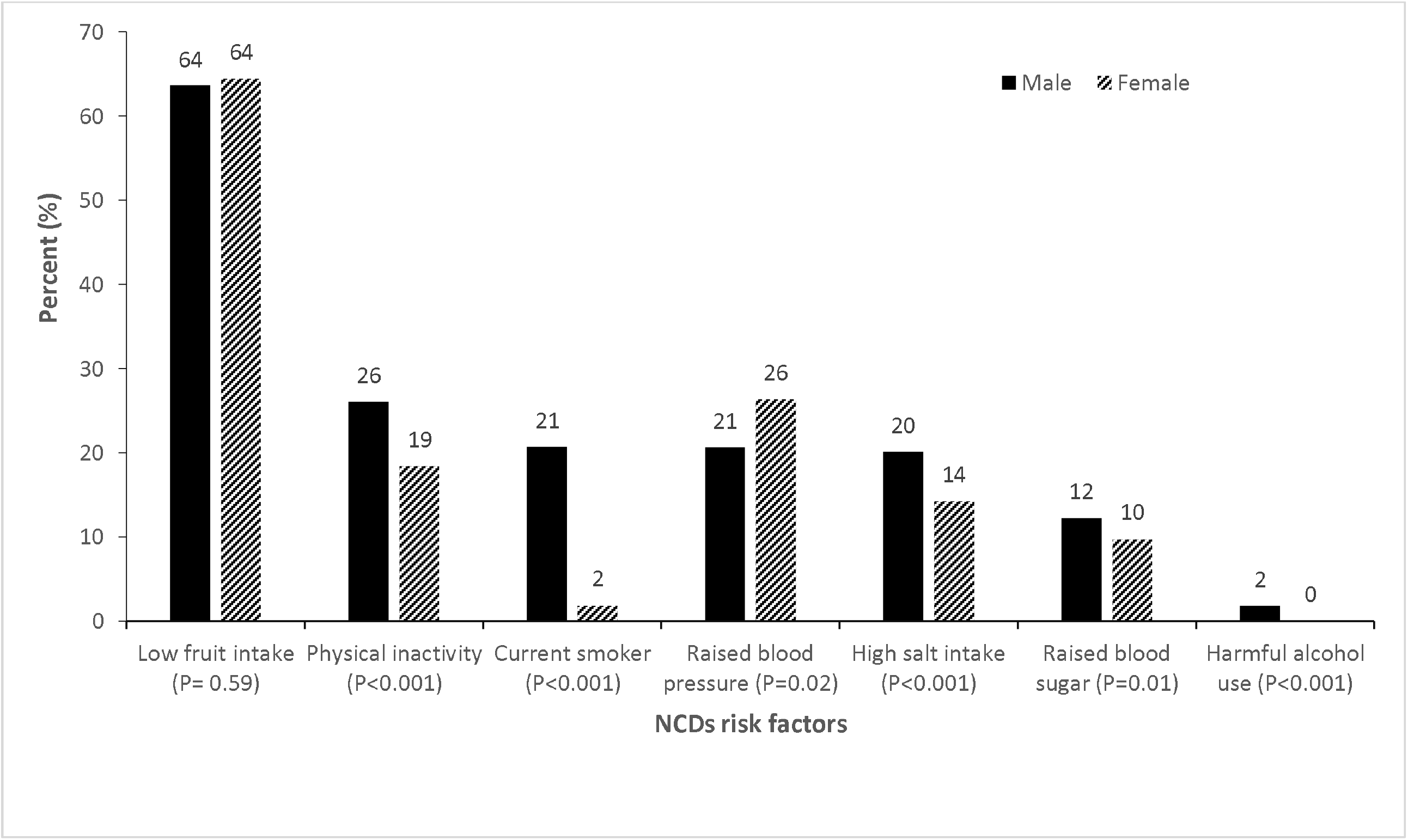
NCD risk factors by sex: 2017 STEPS survey.

#### Harmful alcohol use

The prevalence of harmful alcohol use was 1%. The persons aged between 25 years and 35 years (1%), those with tertiary education (2%), those who were never married (1%) and the self-employed (1%) had the highest harmful alcohol use (see Table 2). More men (2%) than women (0%) indicated that they had harmful alcohol use (see Figure 1).

#### High salt intake

The overall prevalence of high salt intake was 16%. The persons aged between 25 years and 35 years (18%), those with tertiary education (30%), those who were never married (20%) and the government employees (23%) had the highest proportions with high salt intake (see Table 2). More men (20%) than women (14%) had high salt intake (see Figure 1).

#### Low fruit intake

Insufficient fruit intake was prevalent among 64% (95%CI: 63-66). There was a declining trend in the proportion of persons with insufficient fruit use with increasing age (Table 2). The persons who were never married had the highest proportions of those with insufficient fruit uptake (69%) compared to those who were previously married (61%). Low fruit uptake was most prevalent amongst the government employees (see Table 2). There were similar men and women with insufficient fruit uptake (Figure 1).

#### Physical inactivity

Physical inactivity was observed in 21% (95%CI: 20-22) of persons. Physical inactivity reduced with increasing age although it increased with increasing level of education. Physical inactivity was most prevalent amongst those who were never married (32%) and government employees (37%). More men (26%) than women (19%) had physical inactivity (Figure 1).

#### Raised blood sugar

The overall prevalence of persons with raised blood sugar was 25% (95%CI: 23-27). The proportion of persons with raised sugar increased with increasing age (Table 2). On the other hand, the proportion of persons with increased sugar decreased with increasing level of education. Having raised sugar was most prevalent amongst the divorced or widowed (30%) and employees of non-governmental organizations (29%). A similar proportion of men and women had raised blood sugar (Figure 1).

#### Raised blood pressure

The overall prevalence of persons with raised blood pressure was 11%. The proportion of persons with raised blood pressure decreased with increasing age (Table 2). On the other hand, the proportion of persons with increased blood pressure increased with increasing level of education. Having raised blood pressure was most prevalent amongst the those who never married (15%) and employees of non-governmental organizations (14%). More women (26%) than men (21%) had raised blood pressure (see Figure 1).

### Determinants for NCDs risk factors in Malawi

The factors associated with smoking, harmful alcohol use, high salt intake and low fruit intake are shown in Table 3. The factors associated with smoking are education level, marital status, sex and age. The females were less likely to smoke compared to males (AOR=0.05, 95%CI: 0.04-0.07, P<0.001). The likelihood for smoking increased with increasing age, however; the likelihood for smoking decreased with increased level of education (Table 3). In regard to harmful use of alcohol, females had lower odds of engaging in harmful use of alcohol (AOR=0.04, 95%CI: 0.01-0.17, P<0.001). With regard high salt intake, females had lower odds of high salt uptake than the males (AOR=0.70, 95%CI: 0.58-0.84, P=0.0001). The persons with at least secondary education had higher odds of high salt uptake with increasing age (Table 3).

**Table 3:**
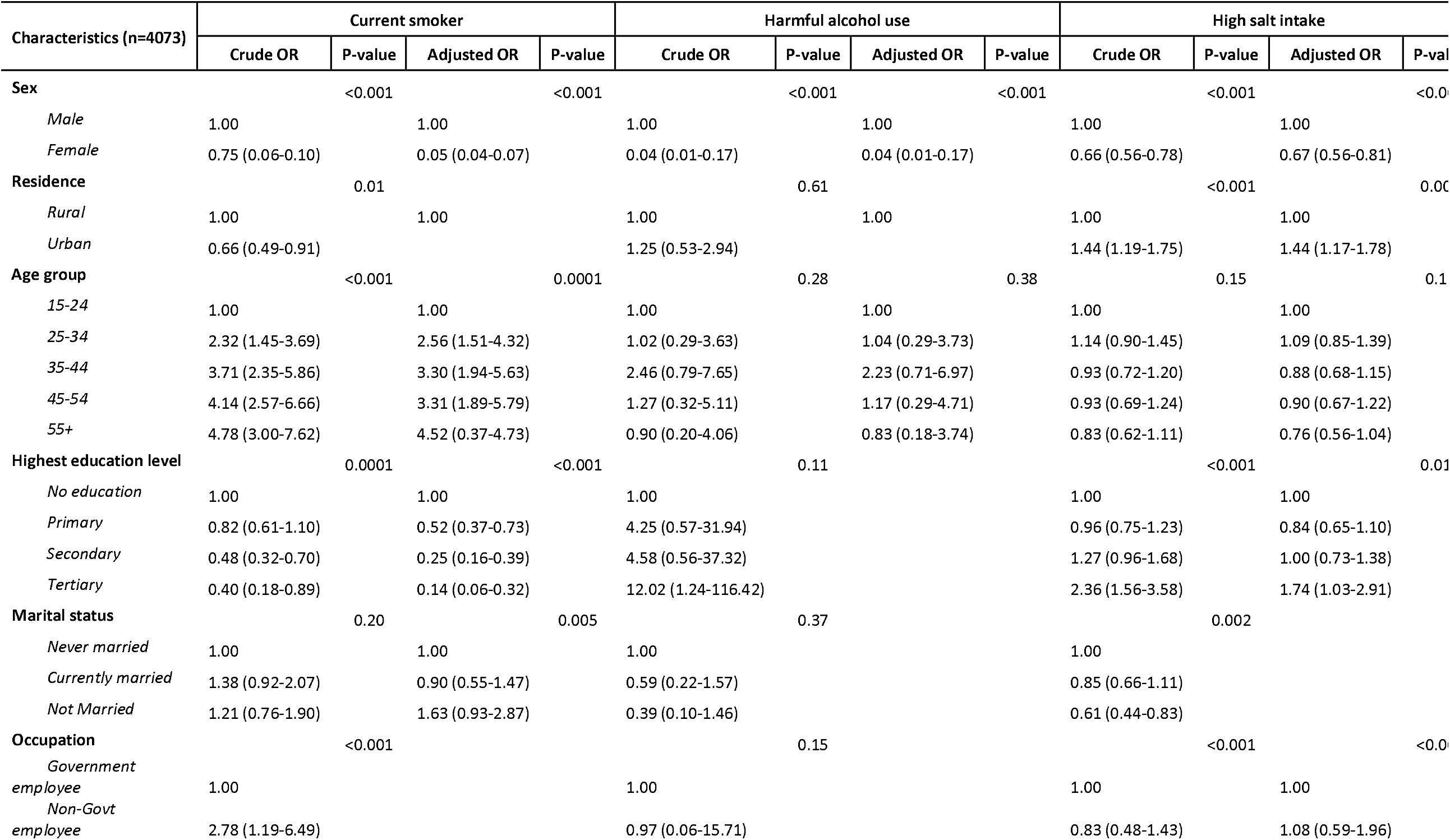
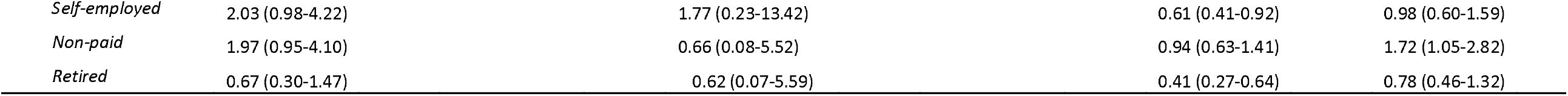
Determinants of selected risk factors of NCDs in Malawi: 2017 STEPS survey.

Persons in non-paid jobs had higher odds of salt uptake than the government employees (AOR=1.70, 95%CI: 1.03-2.79, P=0.04). In regard to fruit intake, women were 22% more likely to have insufficient fruit uptake compared to males (AOR=1.22, 95%CI: 1.06-1.41, P=0.007). There was a declining trend in the proportion of persons with insufficient fruit uptake with increasing age, however; increasing level of education was associated with increasing odds of insufficient fruit uptake (Table 3).

The factors associated with physical inactivity, raised blood sugar and raised blood pressure are shown in Table 4. Women had lesser odds of physical inactivity than the men (AOR= 0.79, 95%CI: 0.67-0.94, P=0.006). There was a decreasing trend in the odds of physical inactivity with increasing age, however; there was an increasing trend in the odds of physical inactivity with increasing level of education (Table 4). The persons of other occupations had lesser odds of physical inactivity than the government employees (Table 4). The odds of raised blood sugar were higher amongst the females compared to the males. Besides, there was an increasing trend in odds of raised blood sugar with increasing age (Table 4). On the other hand, being married or ever been married was associated with lesser odds of raised blood sugar. Although there was an increasing trend in the odds of raised blood pressure with increasing level of education, there were lesser odds in the raised blood pressure with increasing age of the persons (Table 4).

**Table 4:**
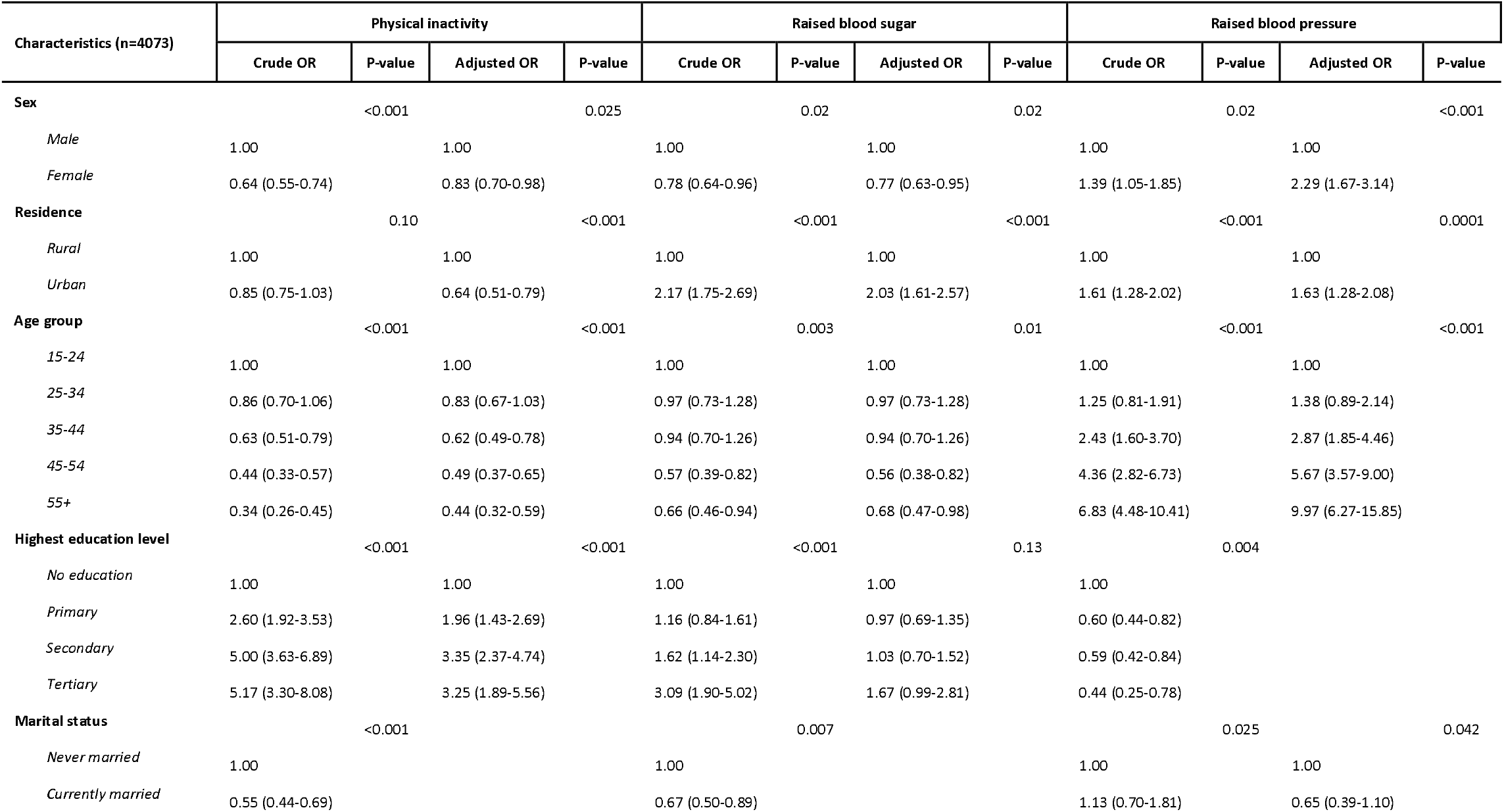
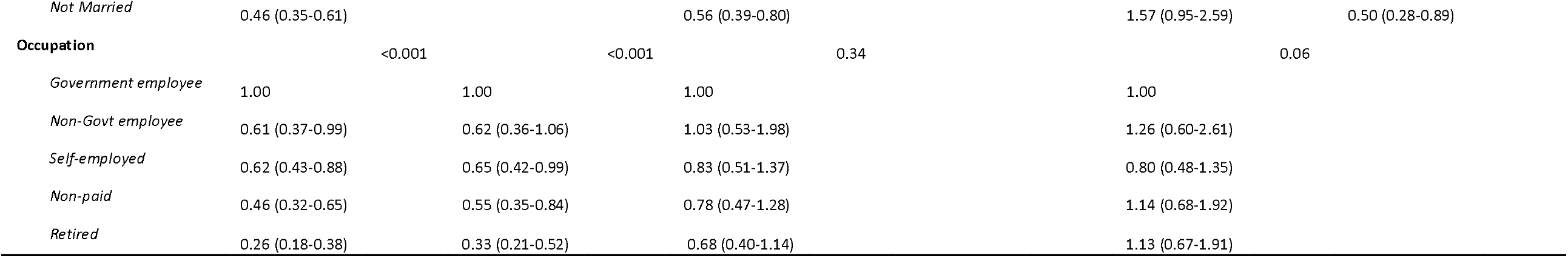
Determinants of selected risk factors of NCDs in Malawi: 2017 STEPS survey.

## DISCUSSION

Our study is the first analysis presenting a comprehensive national overview of the NCD behavioral and biological risk factors for Malawi. The prevalence of tobacco use amongst adult Malawians was not as high as observed in similar settings like Zambia [5], Tanzania, Zimbabwe and Mozambique [13]. In this study and study by Sreeramareddy et. al [13]; there was an increasing trend in tobacco use with age with the older individuals being more likely to use tobacco than their younger counterparts. Furthermore, the males were more likely to smoke tobacco than the females as observed in other similar settings [12] [13] [14]. On the contrary to Sreeramareddy et. al [13], our study shows decreasing trend in smoking with increasing level of education with the observed differences being attributable to different cultural values with regard to smoking in these settings [13]. The prevalence of harmful alcohol use in Malawi was quite low compared to other SSA as shown by Sreeramareddy et. al [13], Boua et al [14] and Tachi et al [15]. In our study, harmful alcohol use was lower in women than men and this is consistent with other studies done in SSA [5] [12] [14] [15] [16].

The salt consumption in Malawi in 2017 was lower than observed in Ghana [12]. There was consistency in rural-urban location as the key determinants of high salt consumption in Malawi [17] and Ghana [12]. This is similar to our study and that conducted by Osei et al [12] and Magali et al [18], and a systematic review which also found that the urban residents had a higher likelihood of high salt consumption [19]. The high uptake of salt in urban is consistent with high likelihood of blood pressure that we observed as there is evidence that high salt consumption is associated with high blood pressure [17]. The high salt uptake amongst the persons with tertiary education may be related to these persons being able to eat roasted or flied meat outside their homes [19] [20]. The high salt consumption amongst the non-paid laborers in Malawi may be attributed to cut on the expenditure for proteins or vegetables eaten together with cereals or starchy foods or they consume a lot of salt to compensate for the lost salt as has been observed in the Middle East [21]. Regulation of salt intake is one of the important factors in controlling high blood pressure, cardiovascular disease, and stroke [12].

Compared to Ghana[12] and Zambia[12], Malawi had a lower proportion of persons with insufficient fruit uptake. The differences may be attributable to the health promotion messages in these settings, differentials in production incentives subsidies and distribution systems, and international trade imbalances [22] [23]. One solution for moving toward a food system in which the availability and accessibility of and need for fruits is by lowering their price and increase their consumption [23]. Similar to a study conducted in the Republic of South Africa [24], those with secondary or tertiary education were more likely to have insufficient fruit consumption. With the high insufficient fruit consumption amongst the females and those in urban areas, there is need for governments in similar settings to have deliberate efforts to target increase fruit uptake amongst their populations.

Osei et al has emphasized the positive effects of physical activity on an individual’s health and well-being [12]. Physical inactivity is associated with development of chronic diseases like type 2 diabetes, cancer, and cardiovascular diseases. Our study found lower proportion of persons that were physically inactive compared to what has been observed in some SSA countries like South Africa [25]. However, Malawi had a higher proportion of persons with physical inactivity than Tanzania [26]. Our findings are consistent with what was observed in Tanzania where the rural residents were more likely to be physically active compared to urban residents [26]. Therefore, sensitizing the urban residents on merits of physical activity may increase the uptake of physical activities and consequently risks as a result of physical inactivity. Contrary to what was observed in Ghana, females in Malawi were more physically active than males [12]. The difference may be attributable to the different gender and cultural orientations in Malawi and Ghana.

Raised blood pressure is a risk factor for numerous chronic diseases like coronary heart disease, chronic kidney disease, and ischaemic heart disease, as well as haemorrhagic stroke [27]. The observed prevalence of persons with hypertension was lower than what has been reported in SSA [28], Ghana [12], Tanzania [29], Malawi [6] and Zambia [5]. Contrary to other studies in the SSA or African region that shows hypertension to be higher with increasing age [6] [28] [29] [30], we observed an decreasing trend in hypertension with increasing age. We also observed a higher likelihood in urban areas and this is consistent with the high salt uptake in the urban areas [31]. The implications of the findings are that regardless of age, the persons should have their blood pressure checked either routinely or during medical care. This could be achieved through integration of blood pressure check-ups in all the health care services being provided in both public and private health facilities in Malawi or similar settings. A population-based survey conducted in Uganda showed consistent findings with our study with the majority of urban residents had type 2 diabetes than the rural residents [32]. The implication of the findings is the need to assess sugar levels in the blood of the individuals to consequently reduce the risk of CVDs at individual as well as population level.

Blood glucose level is used to measure diabetes. In our study, we observed higher prevalence of persons with raised blood sugar compared to Ghana [12] and Zambia [5]. Consistent with study conducted in Ethiopia [31], the prevalence of hyperglycemia varied by age, location and sex. Contrary to the findings from a study in Ghana [12], we found strong evidence of association between age and raised blood glucose. The observed increase in risk of hyperglycemia by age may be attributable to inadequate consumption of fruits and vegetables as well as non-physical activity. The combined effect of age and hyperglycemia has the potential to increase the cases of cardiovascular disease (CVD) in Malawi if life style change interventions are not incorporated at personal or population levels. Compared to a study done in Uganda where the females had lower risk of hyperglycemia, we found that the women had a higher risk of hyperglycemia [32]. The differences may be attributable to differentials in lifestyle as well as dietary behaviours for the two settings. A meta-analysis conducted in SSA also found that hyperglycemia risk was higher in females than in males and this is consistent with most studies done in SSA [33].

The strength of this study is that our findings represent that of the general population of Malawi since this is a national survey. However, the study has some limitations similar to what other studies have reported. The variables assessed in this survey were self-reported, which may have resulted in recall and reporting biases leading to over or underestimation of outcomes [12]. For instance, social desirability bias might lead to underestimation of alcohol use and smoking [12]. With most of the populations aging in the era of HIV/AIDS treatment and care, the 2017 Malawi STEPS did not include HIV or tuberculosis variables despite the existing evidence of the syndemicity of such conditions [34].

## CONCLUSION

In conclusion, this study provides a detailed analysis of some of the modifiable risk factors of NCDs and their socio-demographic determinants in Malawi using the 2017 STEPS. The high prevalence of physical inactivity, high salt consumption, insufficient fruit intake, raised blood glucose and high relatively blood pressure calls for a sound public health approach. The Malawi Ministry of Health should devise a strategy to minimize exposure to risk factors at the population and individual levels so as to reduce immediate, medium and long-term effects. Furthermore, the Malawi Ministry of Health should partner with the Departments, Ministries and Agencies (DMA) dealing with food and agriculture, industry, labour, education, and town planning to plan and implement integrated interventions that minimize the prevalence modifiable risk factors of NCDs [12]. We also recommend more objective means of measurement of NCDs risk factors in future studies. For example, objective measures and tools for capturing salt intake such as sodium urine testing, and glucose and blood pressure measurements could provide a better assessment of exposures as reported by Osei et al [12].

## Data Availability

Available from WHO website

https://www.who.int/teams/noncommunicable-diseases/surveillance/data

## DECLARATIONS

We declare that there is no conflict of interest in publishing this paper.

## AUTHORS’ CONTRIBUTIONS

WFN led the manuscript writing, conducted data management and analysis; JC, TM, MU, JB, FC, DN & JMB advised on the data analysis and policy insights on the paper. All authors read and approved the final manuscript.

## ACKNOWLEDGEMENT

The authors would like to thank the World Health Organisation (WHO) for allowing us to use the Malawi 2017 STEPS data.

